# A functional MRI and magnetoencephalography study of the cognitive modulatory effect of transcranial direct current stimulation in early Alzheimer’s disease

**DOI:** 10.1101/2025.04.02.24314106

**Authors:** Himanshu Joshi, Gowthami Nair, Ashika Anne Roy, Setu Havanur, Subhashini K Rangarajan, Vanteemar S Sreeraj, Preeti Sinha, Mariyapa Narayanan, Keshav Kumar J, Sanjib Sinha, Jitender Saini, Sivakumar P Thangaraju, Mathew Varghese, Ganesan Venkatasubramanian, John P John

**Affiliations:** Multimodal Brain Image Analysis Laboratory, National Institute of Mental Health and Neurosciences, Bengaluru, 560029 India; Translational Psychiatry Laboratory, National Institute of Mental Health and Neurosciences, Bengaluru, 560029 India; School of Liberal Arts and Design Studies, Vidyashilp University, Bengaluru 562110 India; Department of Psychiatry, National Institute of Mental Health and Neurosciences, Bengaluru 560029 India; Department of Neurology, National Institute of Mental Health and Neurosciences, Bengaluru 560029 India; Department of Clinical Psychology, National Institute of Mental Health and Neurosciences, Bengaluru 560029 India; Department of Neuroimaging and Interventional Radiology, National Institute of Mental Health and Neurosciences, Bengaluru 560029 India; Centre for Brain Mapping & ADBS Neuroimaging Centre, National Institute of Mental Health and Neurosciences, Bengaluru 560029 India; Geriatric Clinic and Services, National Institute of Mental Health and Neurosciences, Bengaluru 560029 India

**Keywords:** transcranial Direct Current Stimulation, Alzhieimer’s Disease, Mild Cognitive Impairment, functional Magnetic Resonance Imaging (fMRI), Magnetoencephalography

## Abstract

Anodal transcranial direct current stimulation(tDCS) is known to improve cognition in patients with mild cognitive impairment (MCI) and Alzheimer’s disease(mild AD). We aimed to examine the brain functional alterations accompanying improvement in cognitive performance following anodal tDCS at the left dorsolateral prefrontal cortex(DLPFC) in a sample of patients with early AD (N=40; MCI, n= 19 & mild AD, n=21) using functional magnetic resonance imaging (fMRI) and magnetoencephalography(MEG). The patients showed significant improvements in episodic memory, learning, and delayed recall following tDCS intervention. Significant (p-FDR *<*0.05) reduction in seed (left middle frontal gyrus, lMFG)-to-voxel resting state functional connectivity(rsFC) with precuneus and posterior cingulate gyrus(PCC) was noted following tDCS intervention, while task-based fMRI(tbfMRI) analysis revealed significant (p-FDR *<*0.05) increases in blood oxygen level-dependent(BOLD) activations at PCC and right MFG(rMFG) during episodic memory encoding and retrieval tasks respectively. Furthermore, a significant decrease (p-FDR *<*0.05) in resting state MEG(rsMEG) gamma power at the right occipital cortex and an increase in phase (theta) and amplitude (gamma) coupling at the left entorhinal cortex were observed post-tDCS. The findings of this comprehensive study using resting fMRI and MEG, as well as task-based fMRI, provide mechanistic insights regarding brain functional alterations that underlie the cognitive modulatory effects of anodal tDCS in early AD.

## 1 Introduction

Transcranial direct current stimulation (tDCS) has emerged as a safe, non-invasive brain stimulation (NIBS) procedure for enhancing cognitive functioning in mild cognitive impairment (MCI) and Alzheimer’s disease (AD). ^1,2^ Episodic memory and executive functions are key cognitive domains that are impaired in patients with MCI and mild AD. ^3^ The dorsolateral prefrontal cortex (DLPFC) plays an important role in memory and executive functions^4,5^, and anodal stimulation at the left DLPFC improves accuracy and reaction time during episodic memory performance^6,7^ through modulation of regional electrical activity. ^8^ Furthermore, the cortical excitability of this region is positively associated with working memory performance. ^9^ Anodal tDCS has shown significant promise in enhancing cognitive functions in patients with MCI and AD. ^10,11^ A recent study reported significant improvement in general cognitive functions, immediate and delayed recall as well as new learning ability in patients with MCI, following 10 sessions of home-based anodal tDCS of the left DLFPC.^12^ Thus, anodal tDCS of the left DLPFC can potentially enhance learning, memory, and executive functions. However, there are inconsistencies in the results of studies carried out on limited samples of either MCI or AD. ^2,13–17^

Brain functional alterations accompanying cognitive enhancement following anodal tDCS have been examined using resting and task-based fMRI. However, these studies have generated inconsistent and sometimes conflicting results owing mainly to the differences in the mode of tDCS administered (conventional vs high definition), the site of anodal stimulation, fMRI acquisition type (resting vs task-based), fMRI analysis methods (whole brain, seed-to-voxel, network, and voxel-wise connectivity analyses); as well as sample sizes, which very often were inadequate to derive definitive inferences from whole brain, network-based or whole-brain voxel-wise connectivity analyses. A randomized controlled study in 43 patients with MCI involving the administration of high-definition tDCS (HD-tDCS) over the left DLPFC in the experimental group (n=24) and sham tDCS in the control group (n=19) showed increased intensity of the blood oxygen level dependent (BOLD) signal as measured by the fractional amplitude of low-frequency (0.01-0.1 Hz) fluctuation (fALFF) in temporo-parieto-occipital regions with a bilateral spread, and decreased signal in the right insula, precuneus, superior parietal lobe as well as left thalamus following HD-tDCS. The HD-tDCS group also showed a higher degree of synchronized oscillations measured using regional homogeneity (ReHo), in bilaterally spread fronto-temporal regions and left putamen, though no significant improvement in cognitive performance was observed following the intervention. ^18^ Another study used a double-blind, cross-over, sham-controlled design in 18 patients with MCI that involved resting and task-based fMRI acquisitions to examine semantic word-retrieval performance along with task-related activations and voxel-level functional connectivity using eigenvector centrality mapping (ECM) during administration of anodal tDCS over the left ventral inferior frontal gyrus. Improved cognitive performance during tDCS was associated with reduction in prefrontal hyperactivity noted during the sham tDCS, increase in voxel level connectivity of prefrontal regions and reduced connectivity in more posterior brain regions.^19^ To the best of our knowledge, no previous task-based fMRI (tbfMRI) studies in MCI or AD have reported the effect of anodal tDCS at left DLPFC on BOLD hemodynamic responses in the brain. A sham-controlled study with multi-session anodal tDCS at DLPFC in healthy young adults revealed reduced rsFC within DMN as well as increased rsFC between DMN and bilateral fronto-parietal network (FPN) that accompanied cognitive enhancement. ^8,20^ Recent meta-analyses and systematic reviews of resting and task-based fMRI studies have reported both increased and decreased regional activations and functional connectivity in patients with subjective cognitive decline, MCI, and mild AD^21–23^ as well as following tDCS.^15,24^

Electro/magnetoencephalography (EEG/MEG) gamma power and theta-gamma phase-amplitude coupling (PAC) have been established as markers of cognitive performance^25^ and have the potential to predict disease progression from MCI to AD. ^26,27^ Enhancement of working memory performance following anodal tDCS of the left DLPFC in young, healthy participants was reported to be accompanied by increased theta-gamma PAC.^28^ However, there are no reports of EEG/MEG gamma power or theta-gamma PAC alterations following anodal tDCS in MCI and AD.

Thus, evidence from extant literature for anodal tDCS-induced brain functional alterations that accompany cognitive enhancement in early AD is scanty and inconclusive. The reasons for the inconsistent results may include limited sample sizes in many studies, variability of tDCS (single vs multiple sessions), and the use of different neuropsychological assessment protocols. Moreover, there is a lack of studies that have comprehensively examined such brain functional alterations, combining detailed neurocognitive assessments with resting and task-based fMRI and MEG. We aimed, therefore, to undertake such a study in a sample of patients with early AD (MCI and mild AD). We hypothesized that the modulatory effect of anodal tDCS on cognitive performance in patients with early AD will be associated with alterations of rsFC and tbfMRI activation patterns, as well as enhancement of theta-gamma PAC of resting-state MEG.

## 2 Results

The overall sample of participants with early AD (n=40) (refer Table 1) showed significant improvement in verbal memory scores (Learning Trial 3: Z=-3.586; p *<*0.0001 and Delayed Recall: Z=-3.656 p *<*0.0001) following tDCS (post- vs pre-tDCS). As evident from Supplementary table 1, a substantial number of the items in NNB-E showed trends (p *<*0.05) towards improvement following tDCS, but these differences turned out to be non-significant following correction for multiple comparisons. A significant reduction in seed (lMFG)-to-voxel (S2V) rsFC was noted in bilateral posterior cingulate gyri and precuneus following tDCS intervention (Fig.1). An exploratory post-hoc regression analysis of the S(lMFG)2V-rsFC with neuropsychological performance scores was carried out by correlating the NNB-E subtest scores with the subject-wise rsFC values of both the clusters that showed significant reduction in rsFC after tDCS. This analysis revealed a positive relationship of S2V-rsFC with omissions [cluster size: 223 voxels; r=0.356 (95% CI; 0.041-0.607), t=2.2921 (36), p *<*0.03] and commissions [cluster size: 71 voxels; r=0.349 (95% CI; 0.033-0.601), t=2.2366 (36), p *<*0.03] on the picture cancellation test and a negative relationship of S2V-rsFC with word list learning trial 3 performance [cluster size: 71 voxels; r= −0.374 (95% CI; −0.619-0.061)), t= −2.419 (36), p-value *<* 0.02].

**Figure 1:**
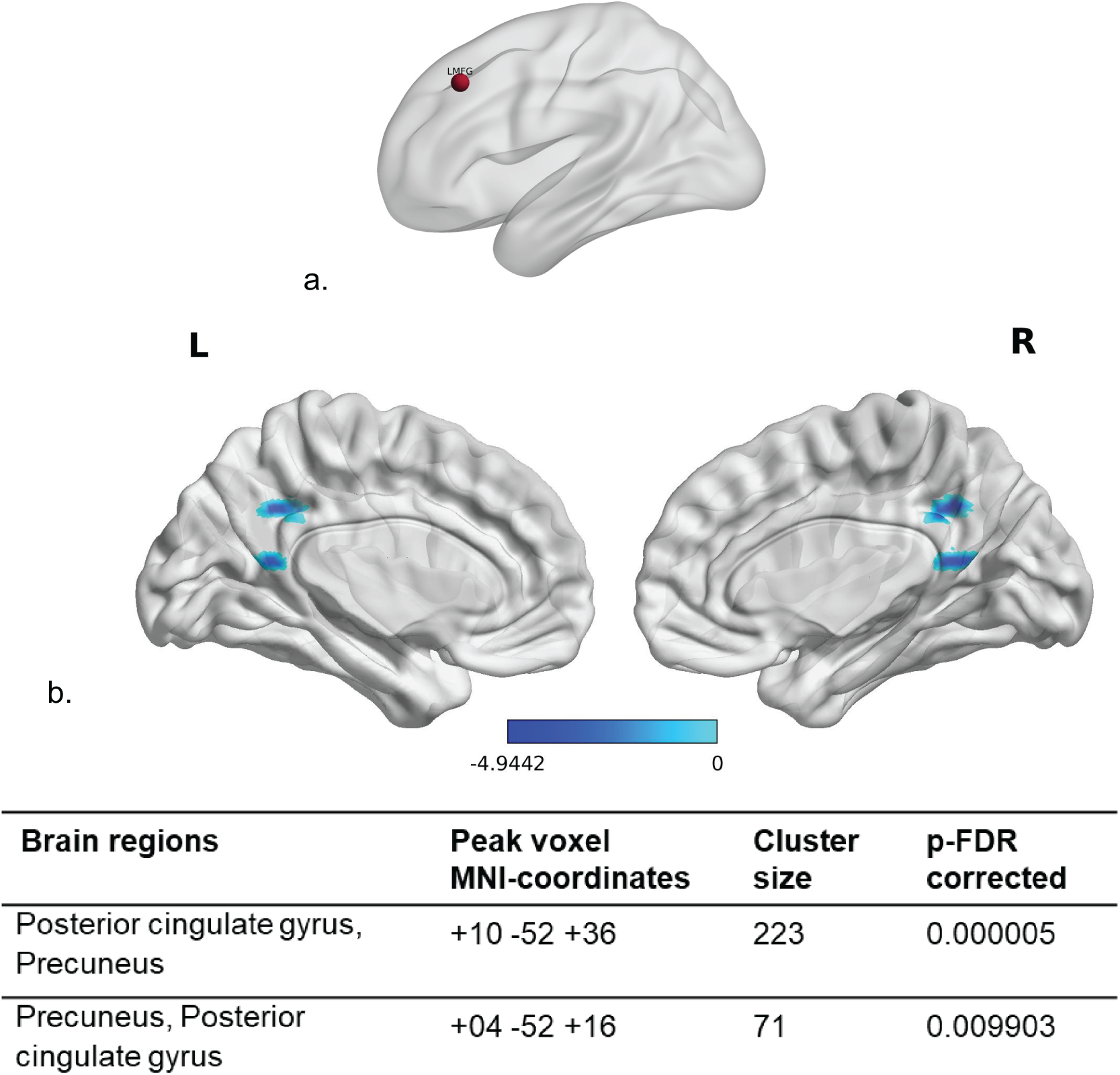
Seed to voxel resting state functional connectivity (S2V-rsFC) analysis: a.) Dark red colored sphere indicates the location of the left middle frontal gyrus (LMFG), which was chosen as the seed for performing S2V-rsFC analysis b.) The light blue to blue color map depicts significantly reduced S(lMFG)2V-rsFC (voxel-wise p-uncorrected *<*0.001 and cluster-wise p-FDR *<*0.05)^29^ at bilateral posterior cingulate gyri and precuneus following tDCS intervention in the early AD [MCI (n=20) + mild AD (n=18)] sample. All the images are coregistered to the MNI template; the cluster coordinates are depicted as per the Conn^30^ functional connectivity toolbox and displayed using the BrainNet Viewer toolbox. ^31^ Refer supplementary figure 4 for original outputs from CONN.

**Table 1:** Socio-demographic and clinical details of study participants.

| Sociodemographic and clinical variable | Overall sample (N=40) | MCI (N=19) | mild AD (N=21) |
| --- | --- | --- | --- |
| Age (years) | 68.37 ± 7.23 | 66.53 ± 6.10 | 70.05 ± 7.89 |
| Education (years) | 13.78 ± 3.52 | 14.68 ± 3.46 | 12.95 ± 3.44 |
| Gender (Male:Female) | 25: 15 | 10: 9 | 15: 6 |
| Number of languages fluent in | 2.90 ± 1.19 | 3.00 ± 1.20 | 2.81 ± 1.21 |
| HMSE1 | 26.13 ± 3.16 | 28.21 ± 2.04 | 24.24 ± 2.79 |
| EASI2 | 3.90 ± 2.59 | 2.10 ± 2.02 | 5.52 ± 1.89 |

MCI: Mild Cognitive Impairment; AD: Alzheimer's disease.
HMSE: Hindi Mental Status Examination.
EASI: Everyday Ability Scale for India.

The analysis of behavioral performance during tbfMRI in the sample of patients who performed at an accuracy level of 60% or greater during both the baseline and post-tDCS fMRI acquisitions (n=30), revealed significant (p*<*0.05) improvement in the accuracy of responses during the incidental encoding task and an overall improvement in the reaction time for both incidental encoding and intentional retrieval trials following tDCS intervention. The discriminability index in the incidental encoding task was noted to improve significantly (p*<*0.05) following tDCS; however, no significant improvement of the discriminability index was noted in the intentional retrieval task (Fig.2). Comparison of BOLD hemodynamic responses between post-tDCS and baseline tbfMRI scans (post-pre) in the above sample of 30 patients (MCI n=15; mild AD n=15) revealed significantly increased activations in the bilateral posterior cingulate gyri during ‘non-living’ trials of the incidental encoding task and increased activations in the right middle frontal gyrus (rMFG) and the right frontal pole (rFP) during ‘non-memory’ trials of the intentional retrieval task using a corrected cluster significance threshold of p*<*0.05^32^ (Fig.2).

**Figure 2:**
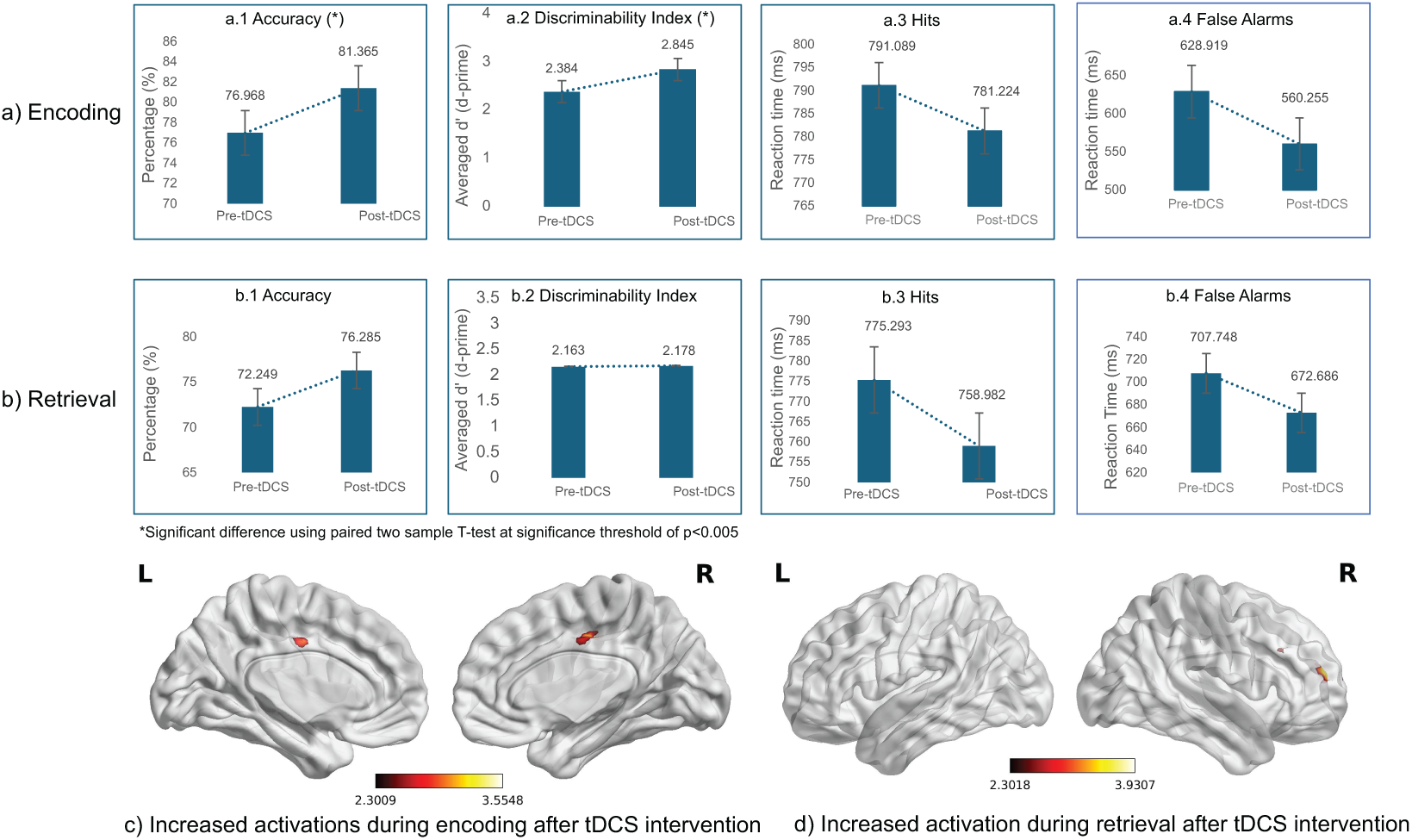
Comparison of performance measures (accuracy, discriminability index, hits, false alarms) during a) encoding and b) retrieval tasks of the episodic memory fMRI paradigm at baseline and following tDCS intervention (pre- vs post-tDCS) in the sample of patients with early AD (N=30; MCI n=15, mild AD n=15) who performed at an accuracy level of 60% or more at both the time points. Significantly increased activations were noted during c) incidental encoding (non-living) trial at the bilateral posterior cingulate gyrus and d) intentional retrieval (non-memory) trial at the right middle frontal gyrus and right frontal pole after tDCS intervention using clusters determined by Z*>*2.3 and a corrected cluster significance threshold of P*<*0.05.^32^ All the activation maps are co-registered to MNI template, the cluster coordinates are depicted according to FSL standards and displayed using BrainNet Viewer toolbox. ^31^ Refer supplementary figures 4 and 5 for original outputs from FSL.

Power spectrum analysis of pre and post-tDCS rsMEG data of patients with early AD (n=34; MCI=15, mild AD=19) showed significantly (FDR corrected p*<*0.05 correcting for multiple comparisons) decreased gamma power in the right occipital cortex after tDCS intervention (Fig.3). The time-resolved phase amplitude coupling (PAC) between rsMEG signals showed that the coupling between the phase of low frequency (theta) and amplitude of high frequency (gamma) increased significantly (FDR corrected p*<*0.05 correcting for multiple comparisons) after tDCS intervention in the left entorhinal cortex (Fig.3).

**Figure 3:**
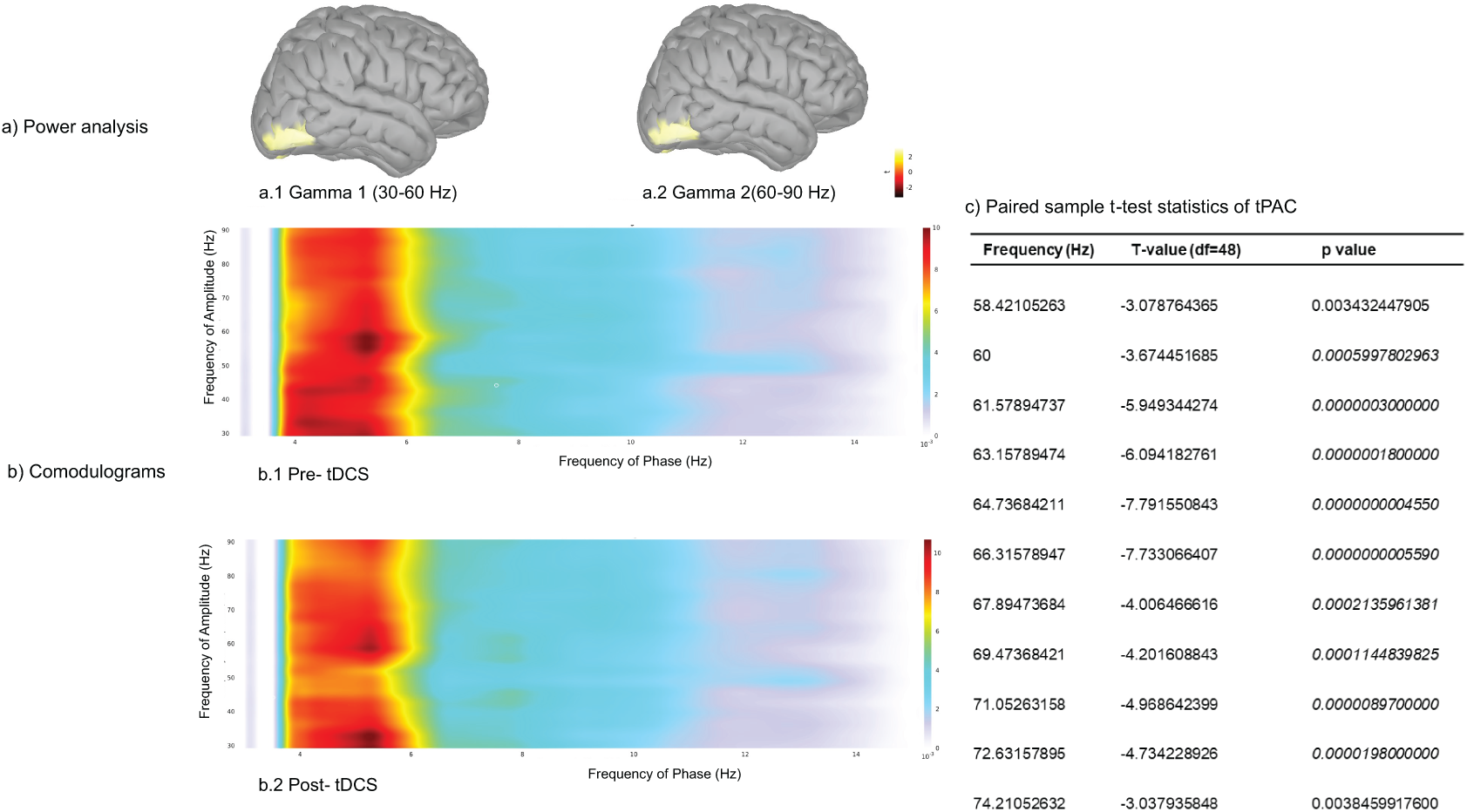
Resting state MEG analyses comparing normalized power spectral density (PSD) (whole brain) and theta-gamma phase amplitude coupling (PAC) (left entorhinal cortex) pre- and post-tDCS in patients with early AD (n=34), correcting for multiple comparisons and controlling for time and frequency. a) Significant PSD reductions (FDR corrected p*<*0.05) of the resting MEG in the higher frequencies a.1) gamma 1 (30-60 Hz) and a.2) gamma2 were noted following tDCS intervention. b) Comparison of average time-resolved PAC (tPAC) comodulograms between pre- and post-tDCS MEG timeseries (b.1 and b.2 respectively) revealed a significant (FDR corrected p*<*0.05) increase following tDCS of PAC between the phase of low frequency oscillations (5.30 Hz) and amplitude of high frequency rhythms (60-72.63 Hz), corrected for multiple comparisons. c) T and p-values corresponding to the those frequencies (60-72.63 Hz) where significant (p= 0.05/39 i.e. p*<*0.00128) increase in tPAC were noted post-tDCS.

## 3 Discussion

The present study aimed to investigate brain functional alterations accompanying the cognitive modulatory effect of anodal tDCS in patients with early AD, using resting-state fMRI and MEG as well as task-based fMRI. We observed significant improvement in verbal memory following anodal tDCS of the left DLPFC in patients with early AD. This was accompanied by a reduction in seed (lMFG)-to-voxel rsFC in the bilateral precuneus and PCC, increased tbfMRI BOLD activations in bilateral PCC and rMFG/rFP during encoding and retrieval tasks, respectively, decreased rsMEG gamma power in the right occipital cortex as well as increased resting theta-gamma PAC in the left entorhinal cortex. These findings provide mechanistic insights regarding the cognition-enhancing effect of anodal tDCS in early AD.

Significant improvement in verbal learning and delayed recall was noted in our early AD sample following anodal tDCS at the left DLPFC. Trend-level improvements were noted in other domains as well, including logical memory, sustained attention, planning, and calculation (vide Supplementary table 1 and fig. 3). In addition, significant improvements were noted in the response accuracy and discriminability index during incidental encoding task performance of the tbfMRI paradigm following tDCS. Trend level improvements were noted post-tDCS in the accuracy and discriminability index during the performance of the intentional retrieval task. The cognitive enhancement noted in the present study, especially in verbal learning, delayed recall, and other domains following anodal tDCS at left DLPFC, replicates the findings of previous studies carried out in patients with MCI and mild AD^2,33–36^ and provides strong support for its potential as an effective intervention for cognition enhancement especially during the early phase of AD. Various mechanisms, including increased dopamine release, ^37^ cortical excitability and neuroplasticity^38^ have been proposed to mediate the above cognition enhancing property of anodal tDCS.

A reduction in S(lMFG)2V-rsFC with bilateral PCC and precuneus was noted in patients with early AD following tDCS intervention. We have previously reported increased rsFC in the executive network in patients with AD^39^ and MCI,^40^ which was interpreted as a compensatory functional response of the relatively preserved neurons to the cortical neuronal loss and white matter disintegration in early AD. ^41,42^ This compensatory hyperconnectivity may be offset by the neuroplastic beneficial effects of tDCS, leading to improved cognitive performance. The PCC is a key component of DMN, which regulates attention, internally directed cognition, spatial memory, configural learning, and retrieval. ^43,44^ Precuneus, another core unit of the DMN, is involved in highly integrated tasks like visuospatial imagery, episodic memory, and retrieval. ^45^ A decrease in PCC-precuneus rsFC was linked in a recent study by Wu et al. ^46^ to improvement in immediate-recall ability in patients with MCI who were trained to enhance their episodic memory. We performed a post-hoc regression analysis of the S2V-rsFC with neuropsychological performance scores to further examine the link between rsFC and cognition, which revealed significant positive correlations between the rsFC values at PCC and precuneus with the omission and commission scores of the picture cancellation task, respectively; and a negative correlation between rsFC values of both the PCC and precuneus with performance on learning trial 3. These findings along with the recent observations by Wu et al. ^46^ provide compelling evidence for decreased rsFC involving bilateral PCC and precuneus as imaging markers of the cognition-enhancing effect of anodal tDCS in early AD.

The tbfMRI results showed increased activations following tDCS intervention in bilateral PCC during the performance of the incidental encoding task; and in the rMFG and rFP during the intentional retrieval task. This may reflect the normalization of brain functioning following anodal tDCS by enhancing the prefrontal cerebral blood flow. ^15,24^ However, the increased activation in bilateral PCC was seen only with the ‘non-living’ trials of the incidental encoding task, while the increased activations in the rMFG and rFP were noted only with the ‘non-memory’ trials of the intentional retrieval task. The findings of this initial study that has used tbfMRI to explore the effect of anodal tDCS on brain activations during episodic memory encoding and retrieval may, therefore, be considered preliminary, and indicate improved activation of brain regions involved in spatial information processing, ^47^ episodic memory and recognition^48–50^ during encoding as well as regions involved in attention, working memory and prospective memory ^51,52^ during retrieval.

The rsMEG results revealed a significant reduction in gamma power in the right occipital cortex following tDCS intervention, along with a substantial increase in theta-gamma PAC in the left entorhinal cortex. This is the first demonstration of altered rsMEG markers of improved cognitive performance^25^ following anodal tDCS intervention in patients with early AD. Though the causal mechanisms by which the gamma rhythms influence specific brain functions remain unclear, it is widely understood that the different bands of gamma oscillations serve an orchestration function to synchronize neuronal populations that are involved in various brain functions in space and time. ^53^ It follows therefore that increased level of synchrony does not necessarily imply a computational advantage, ^53^ and could in fact represent an aberrant pattern of connectivity in pathological states. Increased gamma-band power ^54^ and gamma-band coherence^55^ have been reported in AD and MCI and may thus reflect an effortful compensatory inter-regional integration in the face of the progressively evolving neurodegeneration in early AD. Anodal tDCS has been shown to have strong remote gamma band effects in cortical regions that are located far away from the site of stimulation. ^56^ Thus the reduction of right occipital gamma power in the present study reflects the mitigating effect of anodal tDCS at left DLPFC on the above-mentioned compensatory increase of gamma power in patients with early AD.

The PAC of the rsMEG, an index of how high-frequency oscillatory activity is organized according to the phase of a low-frequency oscillation, is critical for multiple components of episodic memory. ^57^ The results of our study demonstrate for the first time how anodal tDCS intervention significantly increases theta-gamma tPAC in the left entorhinal cortex in early AD, commensurate with significant improvement in episodic memory. Increased cross-regional PAC between the entorhinal cortex and parahippocampal cortices has been linked to successful episodic memory encoding using intracranial EEG. ^58,59^ This finding underlines the significance of crossfrequency coupling (CFC) in cognitive performance^53^ and points toward how the enhancement of CFC in the left entorhinal cortex following anodal tDCS, a downstream effect, leads to improved verbal learning and episodic memory in early AD.

To the best of our knowledge, this is the most comprehensive study to date that has attempted to examine the mechanistic basis of cognitive enhancement following tDCS using multiple modalities of functional brain imaging. The findings of the study highlight a very interesting synergy between the fMRI and MEG results. The improved cognitive performance following anodal tDCS intervention in early AD was accompanied by an increase in theta-gamma tPAC of rsMEG in the left entorhinal cortex, which indicates improvement in CFC essential for cognitive performance, as well as increased tbfMRI BOLD activations of critical brain regions involved in the performance of encoding and retrieval. Furthermore, rsfMRI and rsMEG markers that reflect the compensatory functional responses of the relatively preserved neurons in early AD, namely, increased rsFC and increased gamma power, respectively, were noted to be reversed following anodal tDCS.

Though we report our findings on a sizeable sample of patients with early AD in comparison to many previous studies,^13,16,17,60^ the challenges in carrying out such a comprehensive study on elders with cognitive impairment during the COVID pandemic period meant that we only managed to complete the study in a much more limited sample than was originally planned. This may have impacted our tbfMRI findings, which only gave significant results for the ‘non-living’ trials of the incidental encoding task and the ‘non-memory’ trials of the intentional retrieval task. We recruited our sample based on stringent diagnostic criteria as detailed earlier; however, we did not take into account the more recently suggested biological framework of the NIA-AA for the diagnosis of AD. ^61^ Nonetheless, this is unlikely to have affected our findings, as recently reported by Kang et al. ^62^ in their study on the impact of tDCS on cognition in patients with MCI.

## 4 Methods

This study was conducted at the National Institute of Mental Health and Neurosciences (NIMHANS), Bengaluru, India, with the approval of the NIMHANS Institutional Ethics Committee.

### 4.1 Study Sample

All consenting participants of both sexes (age range: 55 – 84 years) meeting the inclusion and exclusion criteria (see below), who consecutively attended the outpatient services of the Geriatric Clinic and Services (GCS), NIMHANS from August 2019 to December 2022 with memory complaints were recruited in this study with their signed informed consent. Only right-handed individuals were recruited as assessed using the Edinburgh Handedness Inventory (EHI).^63^ Participants were screened to ensure that only those with optimal hearing and visual acuity were recruited. The clinical diagnoses of MCI and mild AD made by clinicians at the GCS at NIMHANS were confirmed using the National Institute on Ageing – Alzheimer’s Association (NIA-AA) diagnostic criteria^64,65^ and Clinical Dementia Rating [CDR]^66^ (CDR scores of 0.5 for MCI and 1 for mild AD). Participants with CDR scores above 1 were excluded from this study. Other exclusion criteria included concurrent medical conditions like hypothyroidism, hypercalcemia, vitamin B12 deficiency, uncontrolled hypertension and diabetes, neurosyphilis, normal pressure hydrocephalus, subdural haematoma, etc., requiring initiation of medical/surgical interventions and/or titration of medications; pre-existing major psychiatric and neurological illnesses, such as schizophrenia spectrum disorders, bipolar affective disorder, major depressive disorder, obsessive-compulsive disorder, substance dependence, intellectual disability disorder, Parkinson’s disease and related disorders, stroke, epilepsy, and other chronic neurological/neurodegenerative disorders, and significant head injury; and current usage of antipsychotics, antidepressants, and benzodiazepines. Forty patients (19 MCI and 21 mild AD) consented to participate in the study and completed the tDCS intervention protocol, which comprised daily 20-minute tDCS sessions for 10 days. All participants underwent rsfMRI and tbfMRI, rsMEG, as well as detailed neuropsychological assessments at baseline and following tDCS intervention.

### 4.2 Clinical and cognitive assessments

The Hindi Mental Status Examination (HMSE), ^67^ a cognitive screening tool validated for the Indian population, and the Everyday Abilities Scale for India (EASI),^68^ a scale validated for the Indian population for assessing activities of daily living, were administered to all participants. Detailed cognitive assessment was conducted using the NIMHANS Neuropsychological Battery for Elderly (NNB-E). ^69^ This battery includes tests of episodic memory, executive functions, attention, visuospatial function, and parietal focal signs and requires 1.5 hours to administer and score. Assessments were carried out at baseline (pre-tDCS) and after completion of the last tDCS session (post-tDCS). These pre- and post-tDCS assessments were carried out by the authors H.J. and S.H., ensuring that the same researcher did not perform both pre- and post-tDCS assessments for a given participant, and the one carrying out the post-tDCS assessment was blind to the pre-assessment scores. Two different forms were used for the pre- and post-tDCS assessments of a participant to avoid practice effects. Task completion times, correct/incorrect answers, and any other significant observations relevant to the assessment were recorded on paper forms. The patients were given adequate rest between assessments as required.

### 4.3 Transcranial direct current stimulation

Transcranial direct current stimulation was administered following standard procedures^70^ using a neuroConn DC-Stimulator Plus device (neuroCare Group GmbH, Munich, Germany) with the anode placed at F3 (left DLPFC) and the cathode over the right supraorbital region (Fp2) using 5×7cm electrodes. The anodal tDCS intervention involved daily sessions (between 10-11 a.m.) for 10 consecutive days, wherein a direct current (DC) of 2mA was administered for 20 minutes (with additional ramp-up and ramp-down phase of 20 seconds each at the beginning and end of the session respectively), adhering to stringent safety measures.^71^ None of the participants reported significant adverse effects.

### 4.4 Magnetic resonance imaging

The MRIs were acquired on a 3Tesla Philips Ingenia CX (Philips Healthcare, Netherlands) scanner with a 32-channel phased-array head coil. Participants were provided with foam pads and earplugs to minimize head motion and noise, respectively. A high-resolution structural MRI sequence was acquired in sagittal orientation with 3-D magnetization prepared rapid acquisition gradient multi-echo (MPRAGE) sequence as per the following protocol: 180 interleaved slices, repetition time [TR] =8 ms, echo time [TE] =3.7 ms, flip angle=80, FOV= 240 mm2, 1 mm3 isometric voxels and acquisition time = 4 minutes and 40 seconds. During rs-fMRI acquisition, the participants were instructed to remain physically and mentally relaxed with their eyes open and not focus on anything particular. For measuring the BOLD signal, an echo-planar imaging sequence (EPI) was acquired using the following parameters: total number of volumes= 275, TR=2000 ms, TE= 20 ms, flip angle=800, FOV= 240 mm2, 3 mm3 isometric voxels and acquisition time = 9 minutes and 24 seconds. Two short EPI sequences were also acquired in both the phase encoding directions to calculate the susceptibility distortion caused by magnetic field in-homogeneities during the EPI acquisition. Quality assurance (QA) of the Philips Ingenia CX scanner, as well as the quality control (QC) of the MRI data, were ensured using the standard protocol followed at the laboratory. ^72^

A novel episodic memory paradigm^73^ was used for tbfMRI acquisition (Supplementary figures 1 and 2). Adapted from the Sternberg item recognition task, ^74^ this paradigm comprises two tasks: an incidental encoding task and an intentional retrieval task. In the incidental encoding task, the participants were instructed on each trial to indicate using a button press whether the presented visual images were of living or non-living objects, while in the intentional retrieval task, the instruction on each trial was to indicate using a button press, whether the presented visual images were seen before (memory images, MI) or not seen before (non-memory images, nMI). Before initiation of the incidental encoding task, the instruction slide showed the following: ‘Press “Index finger” for living image and “Middle finger” for non-living image. Kindly do not press any button for the “+” sign’. Similarly, the instruction slide for the intentional retrieval task showed the following: ‘Press “Index finger” for ‘seen image’ and “Middle finger” for ‘not seen image’. Kindly do not press any button for the “+” sign’. The visual images of living and non-living objects were taken from a pool of 260 images in the Snodgrass and Vanderwart picture collection, ^75^ known for their unambiguous and nameable nature. The optimal number of trials for the incidental encoding task (living objects: 55; non-living objects: 55; fixation cross: 56) and the intentional retrieval task (memory images, MI: 55; non-memory images, nMI: 55; fixation cross: 56) was estimated using the fMRI stimulator to maximize the predictable variance between the three trial classes for each of the two tasks. In the incidental encoding task, 18 of the 55 trials of living images were comprised of 3 images (memory images; MIs) repeated 6 times each while the remaining 37 trials comprised 37 unique images. Similarly, 11 of the 55 trials of non-living images were comprised of 2 MIs repeated 6 and 5 times, respectively, and 44 unique non-living images (vide Supplementary Fig. 1). The intentional retrieval task of the paradigm comprised 55 trials of 5 MIs, each repeated 11 times, and 55 trials of unique non-memory images (nMIs), which were not presented in the incidental encoding task (vide Supplementary Fig. 2). In both tasks, no more than two successive trials of the same class were presented (vide Supplementary flowchart 1). Immediately prior to the scanning session, the participants were trained to respond to the trials by following the instructions projected on the screen prior to each of the two tasks.

### 4.5 Magnetoencephalography

The pre- and post-rsMEG data were acquired using the Elekta Neuromag® TRIUX™ MEG Scanner. The scanner, installed in a magnetically shielded room, consisted of 306 sensors, including 102 magnetometers and 204 orthogonal planar gradiometers. Head digitization was done with a 3D digitizer (Fastrak Polhemus) to obtain the shape, orientation, and known locations of the head, which finally produced the 3D model of the head. Two participants (1 in the MCI sample and 1 in the mild AD sample) opted out of taking part in MEG acquisition, and four participants (3 in the MCI sample and 1 in the mild AD sample) did not participate in post-MEG scans; MEG data of the remaining participants (n=34) were preprocessed (vide Supplementary table 3 for sociodemographic details of these participants). The resting eyes closed MEG recordings were carried out in a sitting position employing the following protocol: sampling rate 2000 Hz, empty room recording=2 minutes, resting-state MEG=15 minutes. EEG data were acquired concurrently to ensure that the participants were alert and relaxed during the recording. Electrocardiogram (ECG), electromyography (EMG), and electrooculogram (EOG) were acquired to remove the heart, muscle, and ocular artifacts, respectively. Quality control (QC) of the MEG data was done using MaxfilterTM (ver 2.2), a built-in software for the ELEKTA/Neuromag system. ^76^ Suppression of interference due to environmental and periodic artifacts was carried out using the spatiotemporal signal space separation (tSSS) method. ^77^

### 4.6 Data pre-processing and analyses

#### 4.6.1 Neuropsychological performance

Shapiro Wilk test was performed to check the normality distribution of the differences between pre- and post-tDCS scores for each sub-test (n=26) of NNB-E.^78^ Accordingly, paired t-tests and Wilcoxon signed rank tests were carried out respectively for normally and non-normally distributed sub-test scores to look for significant effects of tDCS intervention (vide Supplementary table 1). Results that survived the Bonferroni correction for multiple comparisons (p*<*0.001) are reported as significant and others (p*<*0.05) as trends. All the statistical analyses were performed in R version 4.0.2 “Taking Off Again” (2020).^79^

#### 4.6.2 Resting-state fMRI

The acquired MR images in Digital Imaging and Communications in Medicine (DICOM) format were converted to Neuroimaging Informatics Technology Initiative (NIfTI) format using dcm2niix version v1.0.20190902 (https://github.com/rordenlab/dcm2niix). After data anonymization, the header information for each subject was inspected and reoriented to standard format using FMRIB Software Library (FSL).^80,81^ The structural and functional MR images of each subject were visually examined for head motion, structural abnormalities, and scanner or acquisition-related artifacts. The functional images were further processed using DVARS (the spatial standard deviation of successive difference images) in FSL to check for volume-to-volume head movement. ^82^ The rsfMRI data of one participant each from the MCI and mild AD groups showed more than 5% intensity variation in two consecutive volumes indicating poor quality of functional images due to motion artifacts. After the exclusion of both pre- and post-rsfMRI scans of these two participants, the data of the remaining 38 participants (n=18 in the MCI group and n=20 in the mild AD group; vide Supplementary table 2 for sociodemographic details of these participants) were further analyzed for changes in rsFC following tDCS intervention.

Analyses of rsfMRI data were carried out on CONN (version 18b), ^30^ a MATLAB® (https://www.mathworks.com) based software, using statistical parametric mapping (SPM; https://www.fil.ion.ucl.ac.uk/spm/, version 7487) in the background. A pre-processing pipeline for volume-based (with indirect normalization to MNI(2X2X2) space) analyses was implemented, which included susceptibility distortion correction using the field map, with phase and magnitude images generated using TOP-UP^83^ in FSL.^81^ The indirect normalization pipeline involved realignment and unwarp, centering, outlier detection, indirect segmentation, normalization, and smoothing using a Gaussian kernel of FWHM at 6 mm for spatial convolution. The CONN’s default denoising pipeline was performed for subject-motion parameter estimation^84^ and outlier scan identification or scrubbing^82^ using default threshold values (z-normalized global brain activation, movement, and rotation) in the artifact detection toolbox (ART). Using a temporal bandpass filter (0.01 Hz to 0.1 Hz), the data was denoised by regressing the principal components from white matter and CSF, determined by anatomical component-based noise correction (aCompCor), ^85^ the outliers, and the six motion parameters with their first-order derivatives and a linear term regressor.

The seed for the seed-to-voxel resting state functional connectivity (S2V-rsFC) analysis was set using the Harvard cortical atlas, as the left middle frontal gyrus (lMFG), corresponding to the left DLPFC, the site of the anodal tDCS. A spherical mask of size 5 mm diameter was created based on the probabilistic intensity and maximum percentage of finding this brain region (Fig.1). Seed-based connectivity (SBC) maps compute the Fisher-transformed bivariate correlation coefficients between the BOLD time series of seed and each voxel inside the brain. The analysis between pre- and post-tDCS intervention was performed using the spherical mask of the lMFG as ‘seed’ and the whole-brain volume for ‘voxels’. Two-tailed parametric statistics were implemented to obtain the results using a cluster-level extent threshold (p-FDR corrected) with voxel threshold at p*<*0.001 (p-uncorrected) and cluster threshold at p*<*0.05 (p-corrected) (Fig.1; displayed using BrainNet Viewer toolbox; ^31^ vide Supplementary material for original outputs from CONN).

#### 4.6.3 Task-based fMRI

In the incidental encoding task, ‘hits’ were taken as the sum of responses to trials of both living and non-living images, whereas ‘misses’ were trials that the participant missed responding. In the intentional retrieval task, the hits comprised the correct responses for trials of both previously seen and unseen images, with misses noted as trials to which the participant failed to respond. For both encoding and retrieval tasks, the participants were instructed not to press any buttons for the fixation (+) trials. False alarms were calculated as the sum of responses recorded for the fixation trials, as well as incorrect responses to the respective trials in both tasks, while correct rejections were the sum of fixation trials without responses. The probabilities of hits, misses, false alarms, and correct rejections were calculated. The inverse of standard normal cumulative distribution (ISNCD) was calculated with the probability of hits and false alarms using a NORMINV function for the calculated probability. The discriminability index (‘d’) for both tasks was calculated by subtracting the ISNCD of false alarms from the ISNCD values of hits. ^86^ Accuracy in percentage was calculated using equation (1)

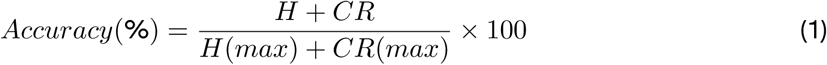

where,

H = Number of Hits
CR = Number of Correct Rejections
H (max) = Maximum number of Hits allowed
CR (max) = Maximum number of Correct Rejections

Three patients with mild AD and five patients with MCI performed with less than 60% accuracy during pre- and/or post-tbfMRI acquisitions; these 8 participants were therefore excluded from further analyses. The performance metrics for the remaining participants (N=30; MCI n=15 and mild AD n=15) for the incidental encoding and intentional retrieval tasks are given in Fig.2. Task-based fMRI data pre-processing and analyses were carried out using FEAT (FMRI Expert Analysis Tool) Version 6.00 of FSL, implementing high-quality model-based fMRI data analysis^80,81^ (Refer to supplementary material for task-based fMRI data preprocessing and analysis). Z (Gaussianised T/F) statistical images were thresholded non-parametrically using clusters determined by Z*>*2.3 and a corrected cluster significance threshold of P=0.05^32^ (Fig.2; displayed using BrainNet Viewer toolbox c; ^31^ vide Supplementary material for original outputs from FSL & Supplementary flowchart 2).

#### 4.6.4 Resting state MEG

The analysis of the available pre- and post-rsMEG data from patients with early AD (n=34; 15 in the MCI sample and 19 in the mild AD sample) was performed. Bad channels (*<* 2% of total channels in any given case) were identified during MEG acquisition and were not considered for further analysis. The rsMEG data was imported using Brainstorm® software for data visualization and manual removal of bad segments. Temporal filtering using a high pass filter (linear phase Finite Impulse Response-Kaiser) of 0.3 Hz to remove DC offset was followed by applying a notch filter up to the third harmonics at 50 Hz with 3-dB bandwidth. Additional bad segments were excluded after visual inspection.^87^ The signal space projection (SSP) technique was used to identify the sensor topographies due to specific artifacts (e.g., blink and cardiac artifact), and its spatial projectors were generated using Principal Component Analysis (PCA) to remove these artifacts.^88^ Additionally, Independent Component Analysis (ICA) was used to filter out the frequency components for eye movement, subject movement, artifacts due to dental implants (between 1-7 Hz), muscle noise, and sensor artifacts (between 40-240 Hz).

For pre-processing the rsMEG data, T1-weighted NIfTI image files from the pre- or post-tDCS MRI sessions with the best QC metrics (see above) were selected. Using FreeSurfer 6.0 software, subject-specific cortical surfaces were reconstructed from these structural MRIs. The pre-processed structural images and the max-filtered MEG data were imported into the Brainstorm® software database, and the subject-specific structural images were used to identify the subject coordinate system. Anterior and posterior commissures and intra-hemispheric points were marked, and each MRI was normalized to MNI coordinates based on the affine co-registration with the MNI ICBM 152 template. Co-registration of the MEG sensors ensured the estimation of source activity with the subject-specific anatomy. Using the overlapping sphere method, forward head modeling was done, followed by source reconstruction using inverse modeling through a linearly constrained minimum variance (LCMV) beamformer advanced source reconstruction algorithm. ^89^ After pre-processing, noise covariance was calculated from empty room recordings to measure the sensor noise, and data covariance was calculated from the resting state MEG recordings.

Using the Welch method, a power spectrum analysis was performed on 5 minutes of artifact-free MEG recording. This analysis estimated the power at different frequencies using a window length of 4 seconds, with a 50% overlap between two consecutive time windows. Extracted power spectrum density (PSD) values for neural oscillations covered canonical frequency bands [delta (2-4 Hz), theta (4-8 Hz), alpha (8-12 Hz), beta (13-30 Hz), gamma (30-60 Hz) and gamma2 (60-90 Hz)]. Spectrum normalization involved dividing the PSD values by the total power of the participant’s source signal that was spatially smoothed with an FWHM of 3 mm. A paired T-test was performed between the pre- and post-tDCS values of normalized PSD, correcting for multiple [permutation testing (1000 randomizations), Monte Carlo method] comparisons (FDR-corrected), controlling for time and frequency.

The dependence between the phase of low-frequency oscillations (fP) and the amplitude of high-frequency rhythms (fA) was quantified using PAC. To estimate the instantaneous phase of the low-frequency filtered signal and the amplitude of the gamma-band signal, the desired spectral range for fP and fA was selected between 4-8 Hz (theta) and 30-90 Hz (gamma), respectively. The coupling strength for each participant was estimated using time-resolved PAC (tPAC), which shows the least relative error. The tPAC was estimated by searching for the fP oscillation with the strongest PAC to fA bursts, over a time window, which slides on the input MEG data. A 10-second epoch length was used for analysis, and since the fP band of interest was between 4-8 Hz, three full cycles of the fP band were chosen to produce a sliding window length of 0.75 seconds. The average comodulograms were extracted using tPAC maps of all the patients with MCI (n=15) and mild AD (n=19) in the left entorhinal cortex^90,91^ which plays a critical role in episodic memory and is vulnerable to early pathophysiology in AD. All the MEG results were generated using Brainstorm® software. ^89^ A paired t-test was performed outside this software using MATLAB® (https://www.mathworks.com) to find out the significant (FDR-corrected p *<*0.05) difference in coupling strength between phase of low frequencies (4-8 Hz) and amplitude of high frequencies (30-90 Hz) correcting for multiple comparisons (n=1911) across time (n=49) and frequency (n=39) (vide Supplementary material for the original outputs and the analysis scripts).

## 5 Conclusion

The findings of this resting fMRI/MEG and tbfMRI study provide mechanistic insights regarding brain functional alterations that underlie the cognitive modulatory effects of anodal tDCS in early AD. The enhancement in episodic memory performance following anodal tDCS of the left DLPFC was noted to be accompanied by alterations of rsMEG and tbfMRI markers that reflect enhanced cross-frequency coupling and regional task-related brain activations respectively, as well as reversal of rsfMRI and rsMEG markers that indicate compensatory functional responses of the relatively preserved neurons in early AD. These have the potential to be considered as markers of treatment response to NIBS procedures in MCI and AD.

## Supporting information

fMRI-MEG-tDCS-Suppl-material

## Acknowledgements

We are indebted to the patients and their families, the healthy participants, as well as the staff members of the ADBS Neuroimaging Centre (ANC) and the Magnetoencephalography (MEG) Centre at NIMHANS for their kind participation in this study.

## Ethics approval and consent to participate

This study was approved by the NIMHANS Institute Ethics Committee [NIMHANS/EC (BEH.SC.DIV.) 14th MEETING/2018 dated 27.10.18]. The privacy rights of all participants in this study have been maintained and informed consent was obtained.

## Data availability

The data used in this study are available from the corresponding author, upon a reasonable request.

## Code availability

Original imaging analysis outputs and the codes used in this manuscript are available at the following link: https://github.com/anshuhim20/pre_post_tDCS_imaging_original_outputs.git

## Author contribution

Authors J.P.J., G.V., and S.P.T have contributed to the conception and design of this study. Authors H.J., A.A.R, S.H. contributed to data collection under the supervision of M.V., J.P.J., G.V., K.K.J, S.P.T, P.S., N.M., and S.S. Authors S.K.R, and H.J. performed the tDCS administration under the supervision of V.S.S., and G.V. Authors H.J. and S.H. administered NIMHANS neuropsychological battery for elderly under the supervision of the K.K.J, who also helped in developing a parallel form for this battery. Authors P.S., S.P.T., M.V., and S.K.R helped recruit participants from the Geriatric Clinic and Services. H.J. performed the analysis and interpretation of results under the supervision of J.P.J., G.V., S.S., and N.M. All the authors suggested revisions to the manuscript critically and approved this version for publication. All the authors agree to be accountable for all aspects of the work in ensuring that questions related to the accuracy or integrity of any part of the work are appropriately investigated and resolved.

## Supplementary information

The supplementary material can be found at fMRI-MEG-tDCS-Suppl-material.pdf

