## Supplementary material for "A functional MRI and magnetoencephalography study of the cognitive modulatory effect of transcranial direct current stimulation in early Alzheimer’s disease": fMRI-MEG-tDCS-Suppl-material

### Incidental encoding (Living/ Non-living discrimination) task:

This is a forced choice stimulus task wherein each stimulus (image) is shown for a duration of 1000 milliseconds, and the inter-stimulus interval (ISI) is varied from 1200-1700 milliseconds with an average ISI of 1500 milliseconds. The right-handed participant is instructed to provide a button-press response to a visual stimulus with the right index finger or right middle finger depending on whether the stimulus is of a living or non-living entity, respectively (Supplementary figure 1). The stimuli presented during this task are expected to be 'incidentally' encoded, in the absence of any *a priori* explicit instructions to hold the images in memory to be utilized while responding to the trials in the subsequent task. The instruction slide prior to the start of the task showed the following: *'Press "Index finger" for living image and "Middle finger" for non-living image. Kindly do not press any button for "+" sign'.*

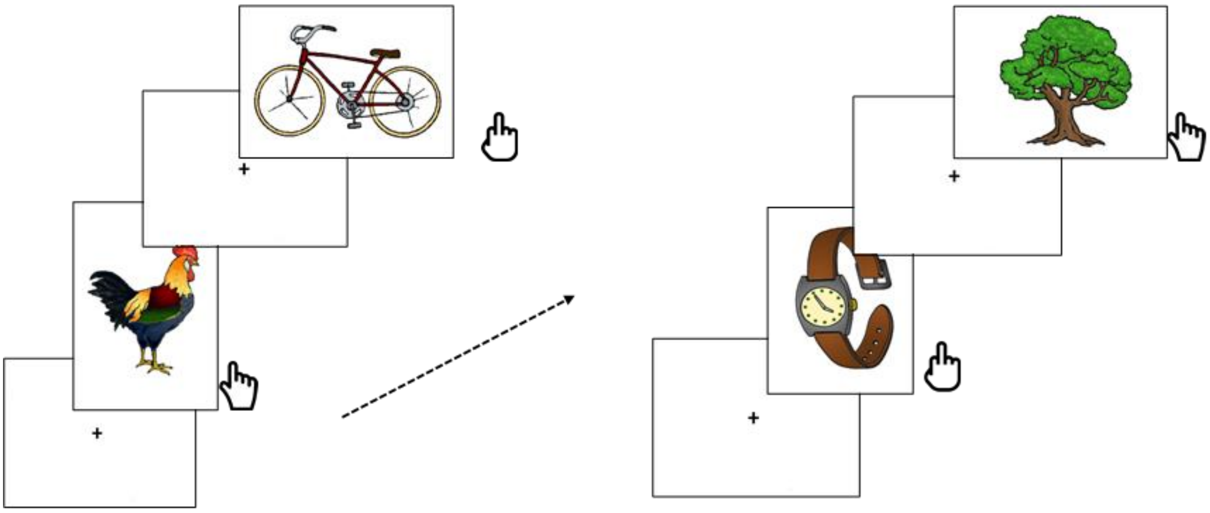

**Supplementary Fig. 1:** A forced choice incidental encoding (Living/ Non-living discrimination) task comprising of 55 trials of living and 55 trials of non-living entities interspersed with 56 fixation trials were presented for a duration of 7 minutes with a restriction that there were no more than 2 sequential presentations of same stimulus type . 3 out of 40 randomly selected living entities were repeated 6 times each, and the remaining 37 images were presented just once. 2 out of 46 randomly selected non-living entities were repeated 6 and 5 times, respectively, with the remaining 44 images presented once. Each trial was presented for a duration of 1000 milliseconds (ms) with randomized jittered inter-trial intervals ranging from 1200 to 1700 (ms) and an average duration of 1500 (ms).

**Intentional retrieval (Memory image/ non-memory image discrimination) task:**

This is a forced choice stimulus task with the same trial parameters as above. In this task, the 5 images which were shown multiple times in the above task are interspersed with new images that were not shown earlier, were presented. The right-handed participant is instructed to provide a button-press response to the stimulus with the right index finger or the right middle finger depending on whether the image was seen before (memory image, MI) or not (non-memory image, nMI) ( Supplementary figure 2). The instruction slide prior to the start of the task showed the following: *'Press "Index finger" for 'seen image' and "Middle finger" for 'not seen image'. Kindly do not press any button for "+" sign'.*

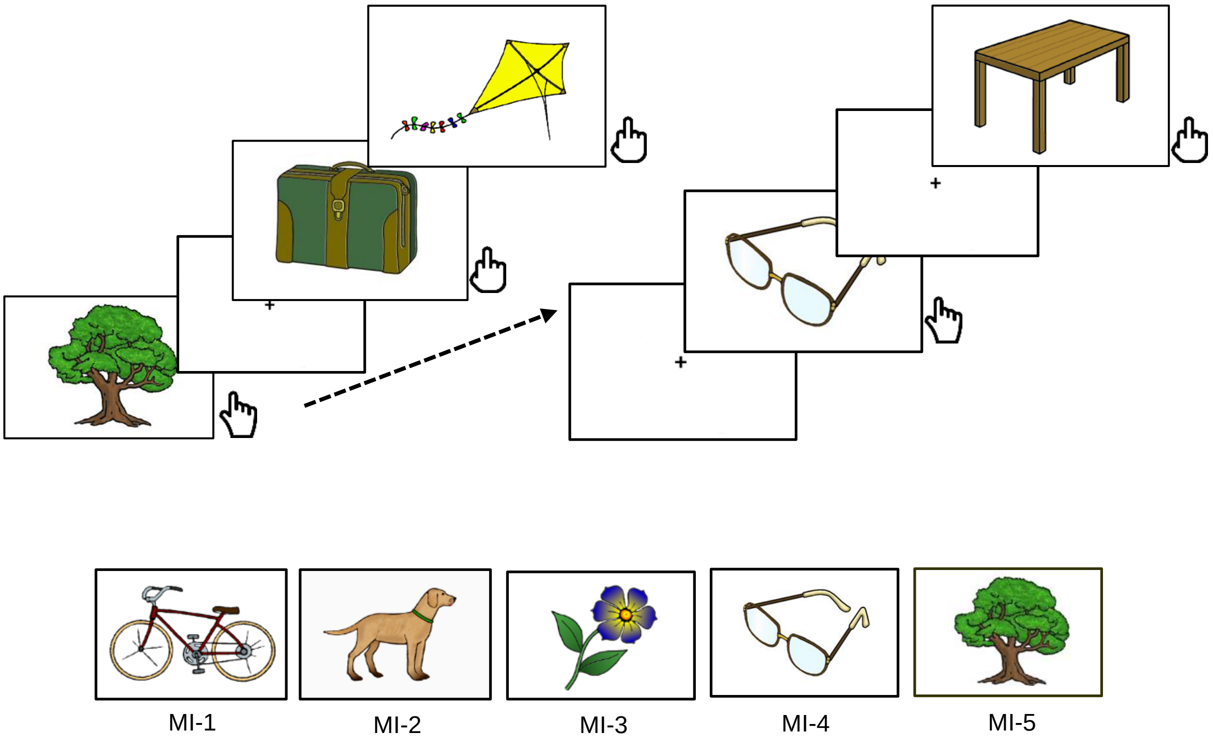


**Supplementary Fig. 2:** A forced choice intentional retrieval (Memory image/ non-memory image discrimination) task comprising of 55 trials of memory images (MI) and 55 trials of non-memory (NMI) images interspersed with 56 fixation trials was presented for a duration of 7 minutes with a restriction that there were no more than 2 sequential presentations of same stimulus type. 5 images repeated 11 times each constituted the 55 memory images, while the 55 non-memory images were unique images not presented before. Each trial was presented for a duration of 1000 milliseconds (ms) with randomized jittered inter-trial intervals ranging from 1200 to 1700 (ms) and an average duration of 1500 (ms).

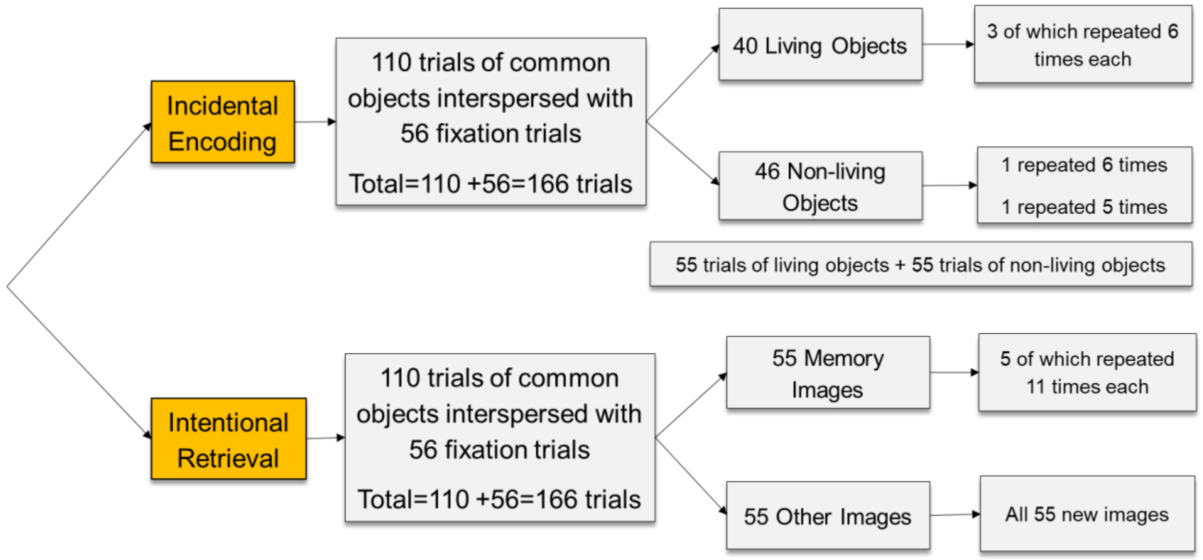


**Supplementary Flowchart 1:** Schematic representation of forced choice incidental encoding (Living/ Non-living discrimination) and intentional retrieval (Memory image/ non-memory image discrimination) task.

### Task-based functional MRI preprocessing and analysis

#### Preprocessing

<u>Distortion Correction</u>

The fMRI acquisition using the gradient-echo EPI sequence was optimized to reduce the signal dropouts during acquisition, while the geometric distortions were corrected by calculating effective EPI echo spacing (0.309 ms) and total readout time (0.0245) as follows:

$$Effective\ Echo\ Spacing = \frac{1}{(Bandwidth\ Per\ Pixel\ Phase\ Encoding\ Direction\ \times Matrix\ Size\ in\ Phase\ Encoding\ Direction)}$$

*Where,*

*Bandwidth-Per-Pixel Phase Encode* = Water-fat shift in Hz / Water-fat shift per pixel

*Water fat shift* in Hz for 3T MRI= 434.215 Hz; Water fat shift per pixel =10.604

The total readout time (according to FSL) is the time from the centre of the first echo to the centre of the last: Also defined as the multiplication of effective echo spacing and the number of reconstructed phase lines.

$$Total\ Readout\ Time\ (FSL) = (Matrix\ Size\ in\ Phase\ Encoding\ Direction - 1) \times Effective\ Echo\ Spacing$$

FMRI acquisition parameters: Matrix Size in Phase Encoding Direction=80; epi factor =39.

Geometric distortion correction was performed in FSLTopUp using two short EPIs with different phase encoding direction and Total Readout Time calculated above to create field map images. These field map images were later converted into the unit rad/s and field map magnitude images were constructed using command “fslmaths” with the option ‘-Tmean’

Motion Correction

Motion correction was carried out to remove the effect of the subject’s head motion during the fMRI experiment using MCFLIRT (Motion Correction FMRIB’s Linear Registration Tool), which uses FLIRT (FMRIB's Linear Registration Tool) by applying rigid-body transformations. Only those patients which showed mean absolute displacement below 2.0 mm and relative rms displacement below 0.5 mm were further analyzed. The residual motion parameter estimates (as estimated by MCFLIRT motion correction described above) were further regressed out.

B0 Unwarping

B0 unwarping was carried out simultaneously with Boundary Based Registration (BBR) to register functional image to structural image[1]. Unwarping was carried out using a fast, automated n-dimensional phase unwarping algorithm. The registration of B0- distorted EPI images was improved using calculated cost function weights[2]. The field map images (constructed earlier), skull-stripped field map magnitude images (reconstructed from the field map sequence), and Effective Echo Spacing of 0.309 milliseconds (calculated earlier) and TE of 20 milliseconds (from EPI data) were used during B0 unwarping in y direction with 10% signal loss threshold implemented in FEAT.

Slice timing Correction

Considering each slice taken at different times within the corresponding volume’s acquisition time (TR), interleaved slice timing correction was carried out using sin c (Hanning-window) interpolation by shifting each time series with an appropriate fraction of a TR relative to the middle of the TR period. Slice timing correction was performed using the Fourier space time-series phase-shifting algorithm.

BET brain extraction

Brain Extraction was applied to Structural data before running FEAT and to functional (EPI) data, using Brain Extraction Toolbox (BET) to improve the quality of registration [3].

Spatial Smoothing

A smoothing kernel of 6mm FWHM was implemented via SUSAN, which provides Gaussian smoothing effectively within the brain while excluding background voxels [4].

Registration

After pre-processing, functional images were registered to the MNI152 standard space (average $T_1$ brain image constructed from 152 normal subjects at the Montreal Neurological Institute, Montreal, QC, Canada) by using affine registration. From the resulting affine transformation matrices, a mid-space was defined as the transformation. This approximates the average size and shape of the individual subjects' spaces by calculating the geometric mean of the affine transformation matrices. Within this mid-space, the data were kept at echo-planar imaging resolutions 3x3x3 mm.

### Analyses

<u>First level</u>

The first level of analysis involved the creation of a data model using the experimental design (based on task-based fMRI paradigm details described above), which should fit the data. Both the task-based experimental (encoding and retrieval) models were created using Custom (3-column format) input waveform in General Linear model (GLM). Each triplet shows a short duration of time and the value of the model during that time: the first number in each triplet shows the onset (in seconds) time, the second number is the duration (in seconds), and the third number shows the value of the input.  A fraction of the blurred original waveform's temporal derivative was added to the model to achieve a slightly better fit to data. FILM (FMRIB's Improved Linear Model)-Voxelwise Timeseries Analysis was performed to non-parametrically estimate the accurate voxelwise temporal autocorrelation to pre-whiten each voxel's time series for the event-related designs with randomized intervals and jittering.

The estimation of noise (percentage of baseline signal) level and temporal smoothness (smoothness coefficient in simple autocorrelation model) was calculated from all the available pre (N=30) and post (N=30) fMRI data. Statistical significance of the level of activation was set to a Z-threshold default value of 5.3 to achieve higher estimates of the required effect. A high pass cut-off value of 90 sec was also estimated from the data, and a filter with the same cut-off was applied to the model. The pre-processing of task-based fMRI data was carried out using the functional (fMRI) data pre-processing steps described above. To remove the residual effects of motion parameters (as estimated by MCFLIRT motion correction described above) that were still left even after motion correction were set as confound explanatory variables (EV). Additionally, the time points corrupted by intermediate to large motions were used as additional confound EVs in GLM to completely remove the effects of these time points on the analysis without having any adverse effects on statistics. This confound matrix was derived using the tool “fsl_motion_outliers”, which performed motion correction, calculated DVARS metric values indicating how much each time point was affected by motion, thresholded the metric values to look for the outliers, and generated the confound matrix for each subject. This format of confound matrix includes the outlier time points with a value of one and all other values as zeros. Adding this matrix to the GLM fully models all the influence of that time point with a separate parameter estimate (PE, or beta); in simple words, intensities at that time point (in any voxel) have no influence on any other parameter estimates. Thus, this model effectively removes the effect of this time point from the estimation of all the effects of interest. This above confound metric was used independently for every subject and were regressed out to calculate voxel-wise pre-whitening matrices using FILM General Linear Model [5]. Double Gamma HRF convolution was applied to the basic waveform to model the standard positive function at normal lag with small late undershoots.

Higher level

Group analysis was performed by implementing FLAME (fMRI Local Analysis of Mixed Effects), the higher-level modelling tool used inside FEAT. To generate generalizability of the results to the wider population, the limited samples of this study were considered as a random quantities with random effect variances[6]. Multi-level (multi-subject and multi-session) modeling was performed using lower-level contrast of parameter estimates derived from the first-level analysis. "FLAME stage 1" model was implemented to estimate the higher-level parameter estimates using a fast approximation to the final estimation. FLAME also implicitly estimated the mixed effects (ME) variance using MH MCMC (Metropolis-Hastings Markov Chain Monte Carlo) sampling, providing the distribution for higher-level contrasts of parameter estimates (COPE). A general t-distribution was then fitted on this distribution for higher-level COPE, and hypothesis testing was then carried out on this fitted t-distribution, providing inference based upon the best estimation of the ME variance [7]. In simple words, a two-stage process (Bayesian modelling and estimation), was used to allow separate modelling of variances in different subject groups and accurately estimated true random effects variance and degree of freedom at each voxel. Automatic outlier de-weighting was also performed for each voxel, considering each subject’s data with respect to other subjects, if it appeared to be an outlier.

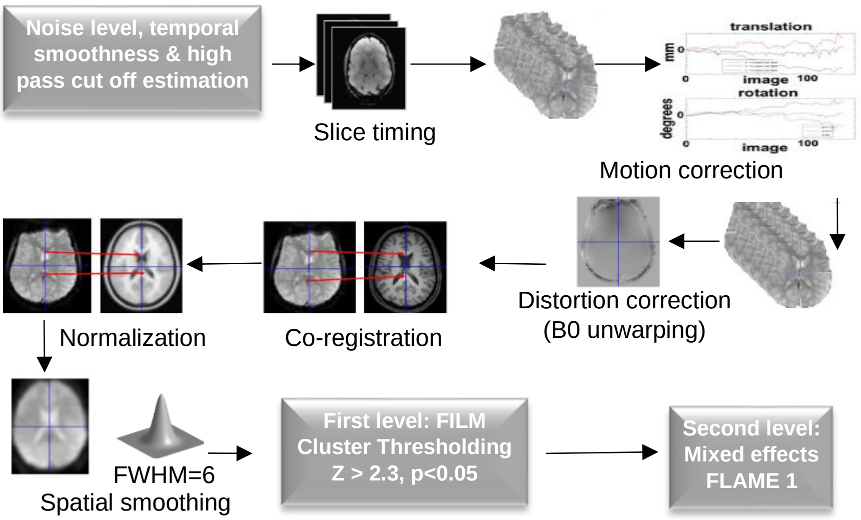


**Supplementary Flowchart 2:** Pre-processing and statistical analysis pipeline for task-based fMRI analysis using FSL

**Supplementary Table 1:** Analysis of neuropsychological assessments of 40 participants MCI (n=19), mild AD (n=21)

| | **Test** | **pre-tDCS** | **post-tDCS** | **Paired t-test result t (df)** |
|---|---|---|---|---|
| **Verbal learning and memory (word list)** | Trial 1 | 3.88(1.20) | 4.20(1.34) | Z=-1883[a], *p*=0.060 |
| | Trial 2 | 5.10(1.53) | 5.48(1.28) | Z=-1.848[a], *p*=0.065 |
| | Trial 3 | 5.85(1.59) | 6.63(1.67) | Z=-3.586[a], ***p<0.0001*** |
| | Delayed recall | 1.50(1.88) | 2.45(2.33) | Z=-3.656[a], ***p<0.0001*** |
| | Recognition (out of 20) | 14.75(3.31) | 14.93(3.71) | Z=-0.388[a], *p*=0.698 |
| **Logical memory (story recall)** | Immediate recall | 6.13(2.70) | 6.83(2.33) | Z=-2.100[a], ***p*=0.036** |
| | Delayed recall | 2.95(3.49) | 3.83(3.51) | Z=-2.677[a], ***p*=0.007** |

| | | | | |
|---|---|---|---|---|
| **Visuospatial learning and memory** | Copy | 23.65(0.83) | 23.78(0.62) | Z=-0.844[a], *p*=0.399 |
| | Immediate recall | 19.33(4.58) | 20.40(4.83) | Z=-1.922[a], *p*=0.055 |
| | Delayed recall | 7.00(6.12) | 7.83(6.54) | t(39)=-1.138, *p*=0.262 |
| **Attention** | Digits forward | 5.15(0.98) | 5.45(1.06) | Z=-2.677[a], **p=0.023** |
| **Working memory** | Digits backward | 3.83(0.84) | 4.08(0.93) | Z=-2.524[a], **p=0.012** |
| **Attention** | Spatial forward | 4.68(0.83) | 5.03(0.89) | Z=-2.462[a], **p=0.014** |
| **Working memory** | Spatial backward | 3.68 (0.86) | 3.93(0.79) | Z=-1.844[a], p=0.065 |
| | Calculations | 7.75 (2.47) | 7.98(3.45) | Z=-2.374[a], **p=0.018** |
| | Fruits | 8.07(3.29) | 8.70(3.30) | t(39)= -1.6591, p=0.105 |
| **Fluency (category)** | Animals | 10.85 (4.15) | 11.63 (4.66) | t(39)=-1.373, p=0.178 |
| | Vegetables | 9.03(4.02) | 9.32(4.18) | t(39)=-0.7076, p=0.483 |
| **Sustained attention (Picture cancellation task)** $ | Time taken to complete (in secs) | 305.16(129.11) | 273.41(98.73) | Z=-3.028[a], **p=0.002** |
| $ One patient couldn't complete this task during pre-tDCS assessment | Commissions | 1.23(2.78) | 0.68(2.31) | Z=-1.765[a], p=0.078 |
| | Omissions | 9.46(10.05) | 6.25(9.95) | Z= -2.612[a], **p=0.009** |
| **Planning (Tower of Hanoi)** $ | Trials completed | 9.26(3.15) | 10.80(2.75) | Z=-3.555[a], **p=0.001** |
| $two patients couldn't complete this task during pre-tDCS assessment | Average time taken to make the first move (in secs) | 5.45(3.62) | 5.25(3.12) | Z=-0.109[b], p=0.913 |

| | | | | |
|---|---|---|---|---|
| | Average moves per trial | 5.98(1.64) | 6.37(1.92) | Z=0.890[b], p=0.373 |
| | Average rule violations | 0.72(0.65) | 0.51(0.60) | t(37)=-2.7136, p=0.010 |
| | Average time taken to solve (in secs) | 27.97(10.84) | 27.20(10.72) | t(37)=0.37, p= 0.711 |

Paired t-test for normal distribution and Wilcoxon signed rank test (a. Based on negative ranks; b. Based on positive ranks) for non-normal distribution after checking post-pre difference scores by the Shapiro-Wilk test.

**Supplementary Table 2:** Socio-demographic and clinical details of patients with early AD (aMCI= 18; mild AD=20) who were included in the fMRI analyses

| **Socio-demographic and clinical variables** | **Early AD (aMCI & mild AD) (N=38)** | | |
|---|---|---|---|
| | *Overall sample (N=38)* | *aMCI (n=18)* | *Mild AD (n=20)* |
| Age (years) | 68.71 ± 7.23 | 66.94 ± 5.99 | 70.63 ± 8.08 |
| Education (years) | 13.50 ± 3.46 | 14.50 ± 3.47 | 12.74 ± 3.41 |
| Gender (Male: Female) | 24: 14 | 10: 8 | 14: 6 |
| Number of Languages fluent in | 2.92 ± 1.21 | 3.06± 1.21 | 2.84± 1.26 |
| HMSE[a] | 26.16 ± 3.18 | 28.22 ± 2.10 | 24.11 ± 2.60 |
| EASI[b] | 3.87 ± 2.65 | 2 ± 2.03 | 5.42 ± 1.89 |

aMCI: amnestic Mild Cognitive Impairment; AD: Alzheimer's disease; HMSE: Hindi Mental Status Examination; EASI: Everyday Ability Scale for India

**Supplementary Table 3:** Socio-demographic and clinical details of patients with early AD (aMCI= 15; mild AD=19) who were included in the MEG analyses

| **Socio-demographic and clinical variables** | **Early AD (aMCI & mild AD) (N=34)** | | |
|---|---|---|---|
| | *Overall sample (N=34)* | *aMCI (n=15)* | *Mild AD (n=19)* |
| Age (years) | 69.09 ± 7.39 | 67.73 ± 6.25 | 70.15 ± 8.20 |
| Education (years) | 13.68 ± 3.51 | 14.93 ± 3.33 | 12.68 ± 3.42 |
| Gender (Male: Female) | 22: 12 | 9: 6 | 13: 6 |
| Number of Languages fluent in | 3.00 ± 1.26 | 3.27± 1.22 | 2.79± 1.27 |
| HMSE[a] | 25.88 ± 3.22 | 28.07± 2.19 | 24.16 ± 2.85 |
| EASI[b] | 4.18 ± 2.58 | 2.33 ± 2.05 | 5.63 ± 1.95 |

aMCI: amnestic Mild Cognitive Impairment; AD: Alzheimer's disease; HMSE: Hindi Mental Status Examination; EASI: Everyday Ability Scale for India

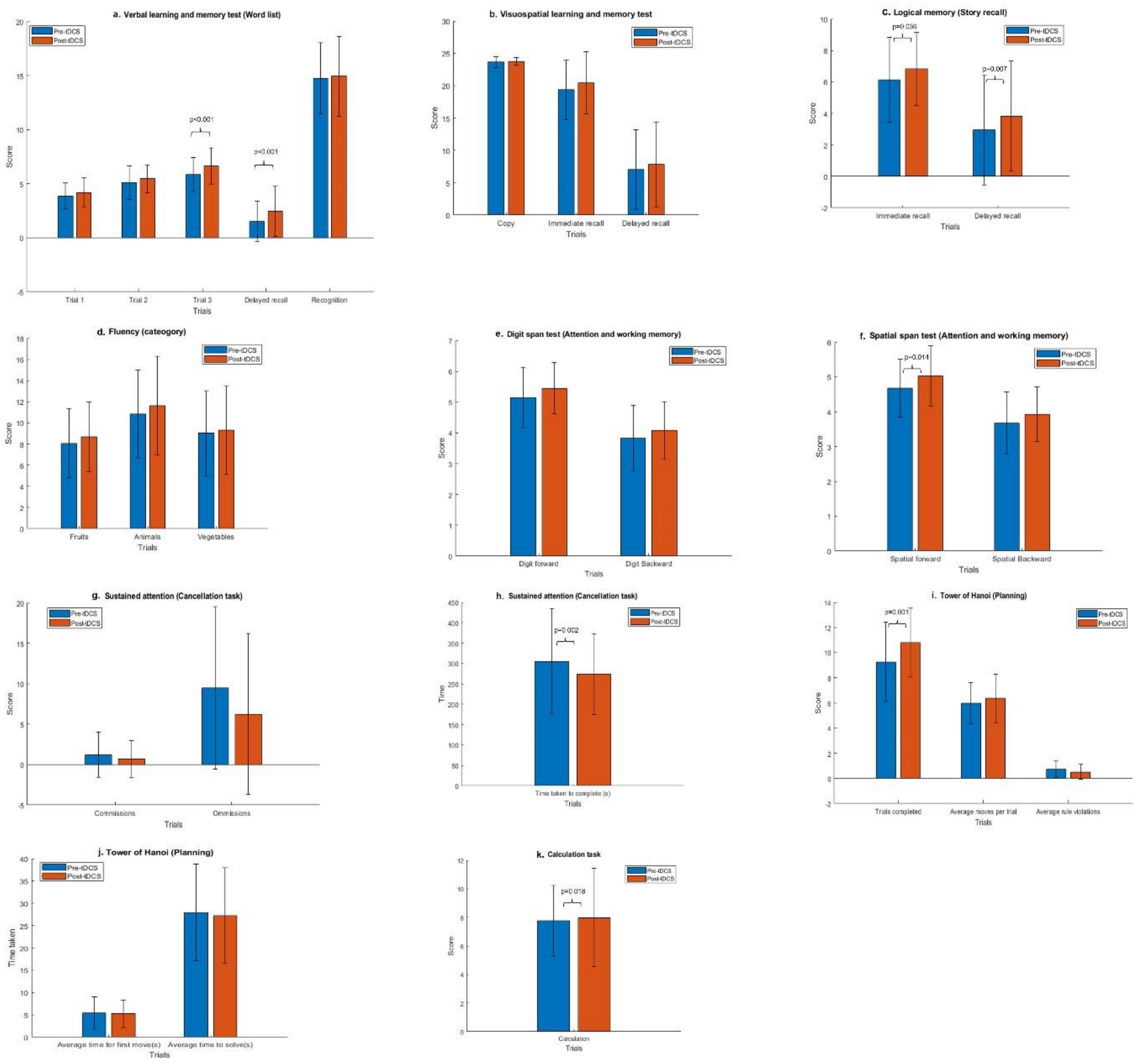


**Supplementary Fig. 3:** Pre (blue)- and post (red)-tDCS cognitive performance scores on a. Verbal learning and memory (Word list), b. Visuospatial learning and memory, c. Logical memory (Story recall), d. Fluency (category), e. Digit span test (Attention and Working memory), f. Spatial span (Attention and Working memory), g. Sustained Attention (Cancellation task), h. Sustained attention (time taken for cancellation), i. Tower of Hanoi (Planning), j. Tower of Hanoi (Time taken for planning) and k. Calculation task from the NIMHANS neuropsychological battery for elderly (NNB-E) in 40 participants with early AD (MCI (n=19), mild AD (n=21)).

**Supplementary Table 4:** Whole brain tbfMRI analysis of incidental encoding (non-living) task showing increased brain activation in the early AD sample (N=30; aMCI *n*=15, mild AD *n*=15), thresholded non-parametrically using clusters determined by Z>2.3 and a (corrected) cluster significance threshold of p-value < 0.05.

| **Brain Structures** | **Voxels** | **Clusters Max** | **Max (x,y,z)** |
|---|---|---|---|
| Cingulate Gyrus*$ (anterior and posterior division), Precentral Gyrus*$ | 452 | 3.55 | 10 -16 40 |

*Structure to which center of mass belongs to $Structure to which cluster belongs to

**Supplementary Table 5:** Whole brain tbfMRI analysis of intentional retrieval (non-memory) task showing increased brain activation in the early AD sample (N=30; aMCI *n*=15, mild AD *n*=15), thresholded non-parametrically using clusters determined by Z>2.3 and a (corrected) cluster significance threshold of p-value < 0.05.

| **Brain Structures** | **Voxels** | **Clusters Max** | **Max (x,y,z)** |
|---|---|---|---|
| Right Frontal Pole*$, Right Middle Frontal Gyrus$ | 564 | 3.93 | 36 54 18 |

*Structure to which center of mass belongs to $Structure to which cluster belongs to

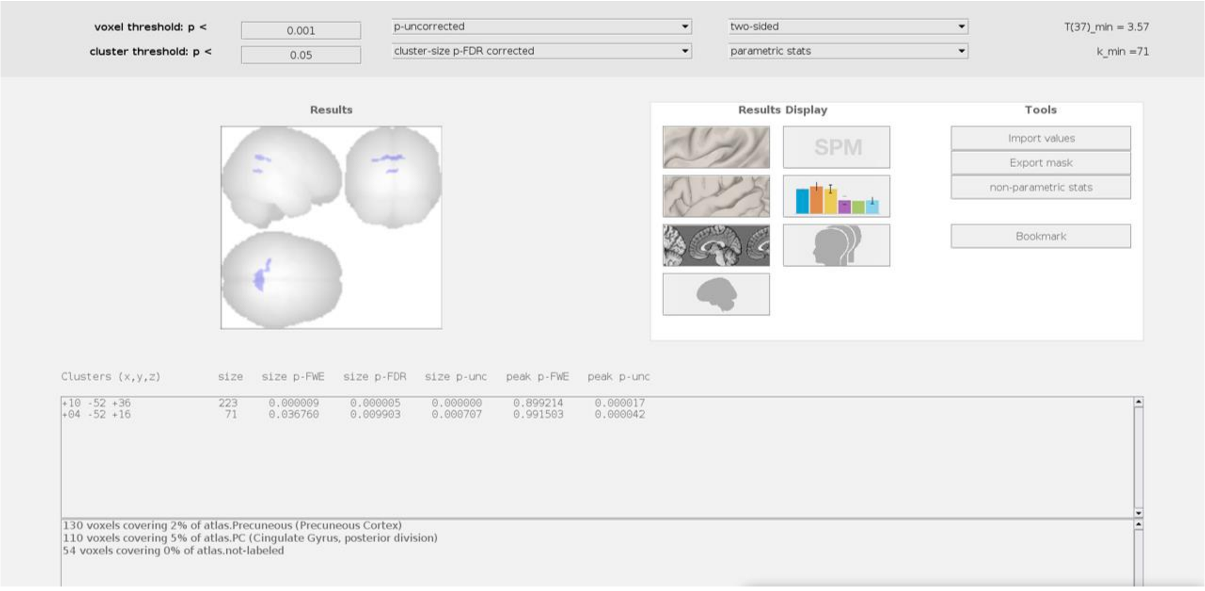


**Supplementary Figure 4.** Original output generated in CONN [8] showing a significant reduction in seed (LMFG)-to-voxel functional connectivity at cingulate gyrus (posterior division) and precuneus after tDCS intervention in the early AD (aMCI (n=18) + mild AD (n=20)) sample using voxel-wise p-uncorrected <0.001and cluster-wise p-FDR<0.05.

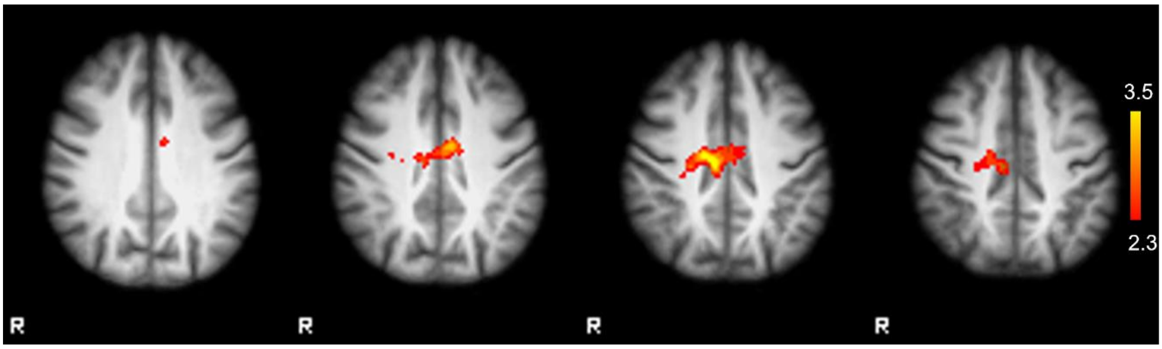


**Supplementary Fig. 5:** Original output generated in FSL [9] showing an increase in activation in right cingulate gyrus (anterior and posterior divisions) for incidental encoding (non-living) trial after tDCS intervention in the early AD (aMCI (n=15) + mild AD (n=15)) sample using clusters determined by Z>2.3 and a (corrected) cluster significance threshold of P<0.05.

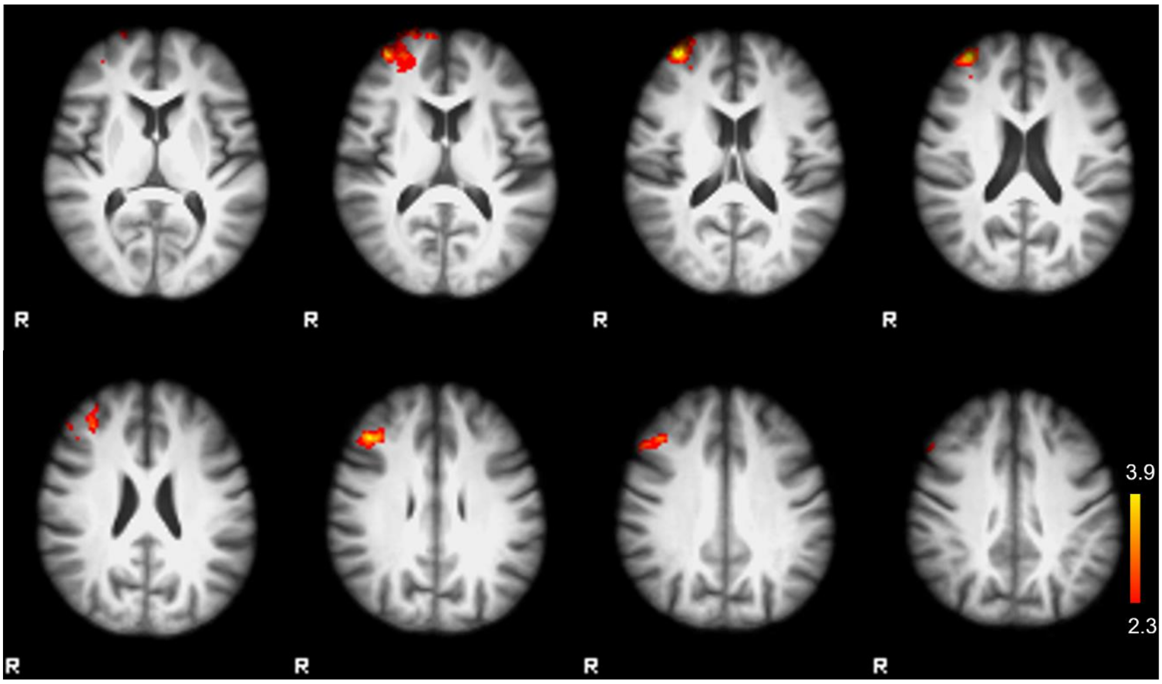


**Supplementary Fig. 6:** Original output generated in FSL [9] showing an increase in activation in the right middle frontal gyrus and right frontal pole for intentional retrieval (non-memory) trial after tDCS intervention in the early AD (aMCI (n=15) + mild AD (n=15)) sample using clusters determined by Z>2.3 and a (corrected) cluster significance threshold of P<0.05.

### Original imaging analysis outputs of this manuscript are available at the following link:

https://github.com/anshuhim20/pre_post_tDCS_imaging_original_outputs.git
